# Towards Internationally standardised humoral Immune Correlates of Protection from SARS-CoV-2 infection and COVID-19 disease

**DOI:** 10.1101/2021.05.21.21257572

**Authors:** Javier Castillo-Olivares, David A. Wells, Matteo Ferrari, Andrew Chan, Peter Smith, Angalee Nadesalingam, Minna Paloniemi, George Carnell, Luis Ohlendorf, Diego Cantoni, Martin Mayora-Neto, Phil Palmer, Paul Tonks, Nigel Temperton, Ralf Wagner, Patrick Neckermann, David Peterhoff, Rainer Doffinger, Sarah Kempster, Ashley Otter, Amanda Semper, Tim Brooks, Mark Page, Anna Albecka, Leo C. James, John Briggs, Wilhelm Schwaeble, Helen Baxendale, Jonathan Heeney

## Abstract

Precision monitoring of antibody responses during the COVID-19 pandemic is increasingly important during large scale vaccine rollout and rise in prevalence of *Severe Acute Respiratory Syndrome-related Coronavirus-2* (SARS-CoV-2) variants of concern (VOC). Equally important is defining Correlates of Protection (CoP) for SARS-CoV-2 infection and COVID-19 disease. Data from epidemiological studies and vaccine trials identified virus neutralising antibodies (Nab) and SARS-CoV-2 antigen-specific (notably RBD, and S) binding antibodies as candidate CoP. In this study, we used the World Health Organisation (WHO) international standard to benchmark neutralising antibody responses and a large panel of binding antibody assays to compare convalescent sera obtained from: a) COVID-19 patients; b) SARS-CoV-2 seropositive healthcare workers (HCW) and c) seronegative HCW. The ultimate aim of this study, was to identify biomarkers of humoral immunity that could be used as candidate CoP in internationally accepted unitage. Whenever suitable, the antibody levels of the samples studied were expressed in International Units (INU) for virus neutralisation assays or International Binding Antibody Units (BAU) for ELISA tests. In this work we used commercial and non-commercial antibody binding assays; a lateral flow test for detection of SARS-CoV-2-specific IgG / IgM; a high throughput multiplexed particle flow cytometry assay for SARS-CoV-2 Spike (S), Nucleocapsid (N) and Receptor Binding Domain (RBD) proteins); a multiplex antigen semi-automated immuno-blotting assay measuring IgM, IgA and IgG; a pseudotyped microneutralisation test (pMN) and electroporation-dependent neutralisation assay (EDNA). Our results indicate that overall, severe COVID-19 patients showed statistically significantly higher levels of SARS-CoV-2-specific neutralising antibodies (average 1029 IU/ml) than those observed in seropositive HCW with mild or asymptomatic infections (379 IU/ml) and that clinical severity scoring, based on WHO guidelines was tightly correlated with neutralisation and RBD / S binding assays. In addition, there was a positive correlation between severity, N-antibody assays and intracellular virus neutralisation.

## 1. Introduction

One year after the World Health Organisation (WHO) declared the COVID-19 epidemic as a Public Health Emergency of International Concern^1^, SARS-CoV-2 has become a global pandemic with more than 3 million deaths recorded up to April 2021^2^. A total of 184 candidate vaccines are now in pre-clinical development and 92 have entered clinical phase. Of the latter, seven vaccines have been approved by National Regulatory Authorities in different parts of the world and WHO have issued Emergency Use Listing for four of these^3^. All these developments occurred in less than a year, thanks to the unprecedented joint effort made by the scientific community, WHO, the pharmaceutical Industry and other philanthropy and international public-health organisations. Because a defined Correlate of Protection (CoP) to COVID-19 did not exist, and still remains elusive, the efficacy of these vaccines was evaluated in large placebo-controlled clinical trials involving large numbers of participants exposed naturally to SARS-CoV-2 in countries that had active COVID-19 epidemics^4^. Though successful, this process was very costly and logistically demanding. The last few months of the pandemic are being characterised by the emergence of SARS-CoV-2 variants that carry mutations that resulted in increased transmissibility, increased pathogenicity or increased potential to evade the immune response of the host^5^. Determining the vaccine efficacy against these variants of concern (VOC) is of high priority for regulatory bodies and vaccine manufacturers in the coming months or perhaps years.

In the absence of a universally accepted CoP against COVID-19, data from Phase III clinical trials suggested that virus neutralising antibodies (nAb) are a candidate CoP ^6^. Likewise, observations made on the natural history of COVID-19 indicated the association of nAb and protection ^7^. However, the protective threshold for nAb has been difficult to establish in different settings. Early studies comparing clinical progression and case fatality rates found that the magnitude of the antibody response correlated with the severity of disease ^8–10^. Patients with fatal outcomes often had amongst the strongest IgG responses to nucleoprotein (N), and were often accompanied by marked responses to the Spike (S) protein ^11^. Furthermore, patients with severe disease have also been reported to have high nAb titres ^12,13^, with studies showing a strong correlation between live-virus or pseudotype based micro-neutralisation (pMN) and anti-Spike antibody binding assays ^10,14^.

Cellular immune responses directed against internal viral antigens often play an important role in the clearance of viral infections. The effector mechanisms of anti-viral immunity of non-neutralising antibodies are becoming better understood ^15,16^ and it is often that these are directed against viral internal antigens, such as the N antigen of SARS-CoV-2. One of these mechanisms is mediated by the cytosolic antibody receptor TRIM21 ^17^, which captures antibody-antigen complexes and accelerates their degradation and processing through the proteasome, facilitating the loading of antigenic peptides in nascent MHC molecules and promoting antigen presentation to T cells ^18^. While this study focused on antibodies against the nucleoprotein of the enveloped positive strand RNA virus LCMV, SARS-CoV-2 anti-N antibodies may be utilized in the same way. We therefore considered TRIM21-mediated biomarkers of immunity in our cohort to determine the holistic role of antibodies in protection from COVID-19 disease.

One of the factors that have precluded the derivation of well-defined humoral CoP in general, and to COVID-19 disease in particular, is the diverse number of quantitative antibody assays available and the different units used to quantify the antibody levels of clinical samples. Measuring nAb against SARS-CoV-2 is typically done by virus neutralisation tests such as plaque reduction neutralisation test (PRNT), infectious centre assays or micro-neutralisation tests ^19–23^. These are performed with live SARS-CoV-2 or with pseudotyped viruses (typically lentivirus or vesicular stomatitis virus) displaying SARS-CoV-2 Spike protein in their envelope. The antibody levels of these assays are quantified in serum titres for specific percentages of neutralisation (i.e. PRNT_50_, PRNT_80_), or as IC_50_ or other readouts. The choice of antibody binding assays is also varied, from the traditional ELISA format to more refined commercial assays (ECLIA, multiplexed micro-sphere assays, semi-automated immunoblotting assays). These methods use the Spike protein or sub-domains thereof or internal nucleoprotein as target antigens. The readouts of these assays are expressed using a diverse suite of units such as antibody titre, OD (optical density) values for specific wave lengths (450 nm, 490 nm) or mean fluorescent intensity (MFI) units or Chemiluminescent units (MCI). In order to harmonise results of quantitative COVID-19 immuno-assays, the WHO has advocated the use of an International Standard SARS-CoV-2 IgG Reagent (20/136), with an assigned unitage, as a primary calibrant ^24,25^. Internal assay calibrants are assigned a potency relative to the primary calibrant which is expressed in International Units. In this way, antibody levels of samples tested by either neutralisation of antibody binding assays can be expressed in International Units and thus immunogenicity and vaccine efficacy data can be compared between different laboratories.

The Humoral Immune Correlates for COVID-19 Project (HICC) aims to dissect the humoral immune response to SARS-CoV-2 and identify the mechanisms of immunity that protect from COVID-19 and to distinguish from pro-inflammatory and complement responses leading to severe disease. The specific aim of the present study is to define antibody-based biomarkers that may serve as humoral candidate CoP and the standardised methods to measure such biomarkers and, where possible, to express them in International Units. Towards this, we have analysed the antibody levels and antigenic specificity of convalescent antibody samples of HCWs and hospitalised COVID-19 patients exposed and infected during the first wave (between March 2020 – October 2020) within the UK. This study defines the methods and findings establishing a benchmark for future longitudinal studies to define COVID-19 CoP.

## 2. Materials and Methods

### 2.1 Selection of sera and plasma

Serum and plasma samples were obtained from HCWs and patients referred to the Royal Papworth Hospital, Cambridge, UK for critical care. COVID-19 patients hospitalised during the first wave and as well as NHS healthcare workers working at the Royal Papworth Hospital in Cambridge, UK served as the exposed HCW cohort (Study approved by Research Ethics Committee Wales, IRAS: 96194 12/WA/0148. Amendment 5). NHS HCW participants from the Royal Papworth Hospital were recruited through staff email over the course of two months (20^th^ April 2020-10^th^ June 2020) as part of a prospective study to establish seroprevalence and immune correlates of protective immunity to SARS-CoV-2. Patients were recruited in convalescence either pre-discharge or at the first post-discharge clinical review. All participants provided written, informed consent prior before enrolment in the study. Sera from NHS HCW and patients were collected between July and September 2020, 3-5 months after they were enrolled in the study.

Clinical assessment and WHO criteria scoring of severity for both patients and NHS HCW (Table 1) was conducted following the ‘COVID-19 Clinical Management: living guidance ^26^.

**Table 1a:** Severity Score Classification.

| Severity Code | Severity Name | Description |
| --- | --- | --- |
| **1** | Asymptomatic |  |
| **2** | Mild Disease | Case definition without of COVID-19 without pneumonia |
| **3** | Moderate pneumonia | Fever, cough, dyspnoea, SpO<sub>2</sub> >90% |
| **4** | Severe pneumonia | Fever, dyspnoea, cough plus RR>30, SpO<sub>2</sub> <90% requirement; |
| **5** | ARDS (Acute Respiratory Distress Syndrome) | diffuse bilateral infiltrates<br>PaO<sub>2</sub>/FiO<sub>2</sub><300 |
| **6** | Sepsis | Life-threatening organ disfunction: severe dyspnoea, delirium, low O<sub>2</sub> saturation, oliguria, tachycardia, weak pulse, low blood pressure, coagulopathy |
| **7** | Septic Shock | As above plus Vasopressor requirement |

**Table 1b:** Cohort Demographic and Severity Score Classification.

|  | Symptom Severity Score |  |  |  |  |  |  |
| --- | --- | --- | --- | --- | --- | --- | --- |
|  | **1** | **2** | **3** | **4** | **5** | **6** | **7** |
| **Patients** | 2 | 3 | 0 | 15 | 1 | 2 | 15 |
| **HCW-P** | 4 | 12 | 8 | 0 | 0 | 0 | 0 |
| **HCW-N** | 22 | 13 | 3 | 0 | 0 | 0 | 0 |

For cross-sectional comparison, representative convalescent serum and plasma samples from seronegative HCWs, seropositive HCW and convalescent PCR-positive COVID-19 patients. The serological screening to classify convalescent HCW as positive or negative was done according to the results provided by a UKAS-accredited Luminex assay detecting N-, RBD- and S-specific IgG, a lateral flow diagnostic test (IgG/IgM) and an Electro-chemiluminescence assay (ECLIA) detecting N- and S-specific IgG. Any sample that produced a positive result by any of these assays was classified as positive. The clinical signs of the individuals from which the sample was obtained ranged from 0 to 7 according using the WHO classification described above. Thus, the panel of convalescent serum samples (3 months post-infection) were grouped in three categories: a) Patients (n=38); b) Seropositive HCW (n=24 samples); and c) Seronegative HCW (n=39) (Table 2).

**Table 2:** Cohorts. Classification of participants according to the serological screening of the sera by the ECLIA, multiplex Micros-sphere, and lateral flow assays.

|  |  | Patients |  |  | Seropositive Staff |  |  | Seronegative Staff |  |  |
| --- | --- | --- | --- | --- | --- | --- | --- | --- | --- | --- |
| **Assay Platform** | **Antigen /Isotype** | Pos. | Neg. | ND | Pos. | Neg. | ND | Pos. | Neg. | ND |
| **Luminex** | N | 36 | 2 | 0 | 22 | 2 | 0 | 0 | 38 | 1 |
|  | S | 36 | 2 | 0 | 19 | 5 | 0 | 0 | 38 | 1 |
|  | RBD | 36 | 2 | 0 | 18 | 6 | 0 | 0 | 38 | 1 |
| **Roche** | S | 34 | 2 | 2 | 18 | 6 | 0 | 0 | 36 | 3 |
|  | N | 34 | 2 | 2 | 18 | 6 | 0 | 0 | 36 | 3 |
| **Lateral Flow** | IgG | 35 | 2 | 1 | 20 | 4 | 0 | 0 | 39 | 0 |
|  | IgM | 19 | 15 | 1 | 15 | 9 | 0 | 0 | 39 | 0 |

### 2.2. Internal and External Calibration Sera

The reference antisera used as external, or primary, calibrants in our assays included: a) the First WHO International Standard for anti-SARS-CoV-2 immunoglobulin (NIBSC 20/136); b) the Anti-SARS-CoV-2 Antibody Diagnostic Calibrant (NIBSC 20/162; and c) the Research Reagent for anti-SARS-CoV-2 Ab (NIBSC 20/130). Details of these are described in the NIBSC catalogue ^27^. We used these external reference reagents to calculate the unitage of tested samples and / or to calibrate our own Internal (or secondary) assay calibrants. The latter were obtained from NHS healthcare workers exposed to SARS-CoV-2. Thus, HICC Serum-1 and HICC Serum-2 were pooled serum samples collected from RT-PCR-confirmed SARS-CoV-2-infected NHS personnel 2 months after presenting moderate symptoms of COVID-19.

### 2.3. Pre-pandemic sera

A panel of 23 pre-pandemic sera collected between 2016 and 2019, obtained from the National Institute of Biological Standards and Control (NIBSC), was used to set up the negative cut-off point of the quantitative immunoblotting assay, a pan-Ig N- and RBD-ELISA and the pMN assays.

### 2.4. Detection of Total Antibody (Pan-Ig) against SARS-CoV-2 Spike (S) and Nucleocapsid (N) antigens by ELISA

Two different ELISA tests were used for the detection of N-specific and S-specific antibodies. The assays were adapted from those originally described by Amanat and co-workers ^28^. Briefly, Nunc MaxiSorp™ flat-bottom plates were coated with 50 μl per well of 1 μg/ml of either RBD or N antigen in DPSB (-Ca^2+^/-Mg^2+^)and incubated overnight at 4°C. The next day, the plates were blocked with 3% milk in PBST (0.1% w/v Tween20 in PBS) for 1 hour. After removing the blocking buffer, 50 μl/well of serum samples, diluted in PBST-NFM (1% w/w non-fat milk in PBST) were added to the plates and incubated on a plate shaker for two hours at 20°C. The plates were washed three times with 200 μl of PBST, and 50 μl of HRP-conjugated goat anti-human Ig (H and L chains) (Jackson ImmunoResearch) was added to each well and left to incubate for one hour on a plate shaker for 1 hour. Plates were washed three times with 200 μl of PBST and 50 μl / well of 1-Step Ultra TMB chromogenic substrate (Sigma) (were added to the plates and the chemical reaction was stopped three minutes later with 50 μl 2N H_2_SO_4_. The optical density at a wavelength of 450 nm (OD450) was measured using a Biorad microplate reader.

All test runs included, in addition to the test sample dilutions, an internal calibrant dilution series (HICC Serum 2), a single dilution of a positive control per plate (NIBSC 20/130), a negative control sample (NIBSC 15/288) per plate and a blank control (no primary antibody or sample). All samples were tested in duplicate and the duplicate readings were used to fit the curve. The blank readings were subtracted from the serum sample values. The IC_50_ values of each sample dilution series were determined and expressed as relative potency respect to the Internal Calibrant, for which a unitage in ELISA binding units was calculated using the WHO International Standard 20/136 as a reference. Details of how these were calculated are described in the ‘Results’ section.

### 2.5. Roche Elecsys® electrochemiluminescence immunoassay (ECLIA)

Samples were tested on Roche cobas® e801 analyser at PHE Porton Down. Anti-nucleocapsid protein antibodies were detected using the qualitative Roche Elecsys® Anti-SARS-CoV-2 (ACOV2) ECLIA (Product code: 09203079190), whilst anti-RBD antibodies were detected using the quantitative Roche Elecsys® Anti-SARS-CoV-2 S (ACOV2 S) ECLIA (Product code 092892751902), as previously described ^29,30^. Both assays detect total antibodies (IgG, IgA and IgM). All kits were calibrated based on a two-point calibration curve according to the manufacturer’s instructions, with daily QC performed per reagent pack. Anti-NP results are expressed as a cut-off index (COI) value, with a COI ≥1 interpreted as positive. Anti-spike results are expressed as units per ml (U/ml), with results of ≥ 0.8 U/ml interpreted as positive and a quantitative range of 0.4 to 2,500 U/ml.

### 2.6. Detection of SARS-CoV-2-S, -RBD and -N specific antibodies using a multiplex bead flow cytometry platform, Luminex™ platform

Detection of serum IgG reactive to SARS-CoV-2 N, S and RBD (receptor binding domain) antigens was done using a Luminex based assay following the methods previously described ^31,32^. The amino acid sequences used derived from the S ectodomain derived from the BetaCoV/Wuhan/WIV04/2019 sequence. All samples were tested in duplicate and all test runs included a serum positive control, a serum negative control, and BSA and LPS antigen controls as blanks. Results outputs were expressed in MFI units. A machine training algorithm was used to assign a final serological classification to all the samples studied, as described previously ^32^. This method assigns a SARS-CoV-2 serological status considering the values the IgG values (MFI) for the three antigens. The negative cut off values for N-, RBD- and S-specific IgG assays were set up at 1604, 456 and 1896 respectively.

### 2.7. SARS-CoV-2 Pseudotype-based microneutralisation assay (pMN)

Virus neutralising antibodies in serum were detected and quantified by a pseudotype-based neutralisation assay based on a lentiviral system that enables the generation of replication-defective recombinant human immunodeficiency virus (HIV) displaying the Spike protein of SARS-CoV-2 on their viral envelope, as previously described ^33,34^. Briefly, HEK293T cells were seeded in 10 cm^2^ cell culture dishes at a density to achieve 70% confluency after 24 hours for next day transfection. HEK293T cells were maintained in DMEM containing 10% foetal bovine serum and 1% penicillin/streptomycin, at 37° C and 5% CO_2_. Cell maintenance was done by three cell passages per week.

On the day of transfection, the culture medium was replaced with fresh complete DMEM. Cells were transfected with 1000 ng of pcDNA-SARS-CoV-2 Spike plasmid, 1000ng of HIV 8.91 gag/pol plasmid and 1500ng of pCSFLW luciferase plasmid, using FuGENE HD (Promega, UK,) at a 1:3 ratio (plasmid:FuGENE HD). The culture media was harvested 48 hours post-transfection and filtered through a 0.45µm filter. The filtered pseudotype virus (PV) was aliquoted, titrated and stored at - 80°C. Titration of PVs was carried out in a 96 well white plate typically using doubling serial dilutions. Pre-transfected HEK293T target cells expressing human ACE2 and TMPRSS2 were seeded at 10^4^ cells per well and plates were incubated for 48 hours prior to the addition of Bright-Glo reagent (Promega, UK) and reading the result in a luminometer.

For detecting and quantifying neutralising antibodies, serial doubling dilutions of the serum samples in coomplte DMEM were performed from an initial 1/40 dilution. SARS-CoV-2 PVs were added at 5×10^5^ – 5×10^6^ RLU/ml in each well and the plates incubated for 1 hour in at 37°C, 5% CO_2_ incubator. Post incubation, pre-transfected HEK293T target cells expressing human ACE2 and TMPRSS2 were seeded at 10^4^ cells per well and plates were incubated for 48 hours prior to the addition of Bright-Glo reagent and assaying using a luminometer.

In addition to the test sample dilutions, all test runs included dilution series of a primary calibrant (NIBSC 20/162) or an internal calibrant (HICC Serum 2) and a single dilution of a positive control per plate (NIBSC 20/130). All samples were tested in duplicate and the average of the OD values determined. The IC_50_ values of each sample dilution series were determined and expressed as relative potency respect to the Internal or external Calibrant which enabled the expression of results in International Units using the WHO International Standard 20/136 as a primary reference. Details of how these were calculated are described in the ‘Results’ section.

### 2.8. Semi-Automated Immunoblotting

Plasma IgG antibodies reactive against the SARS-CoV-2 Spike and Nucleocapsid proteins were analysed by immuno-blotting using the ‘Jess’ fully automated system (ProteinSimple; Bio-Techne) and the SARS-CoV-2 Multi-Antigen Serology Module (ProteinSimple; Bio-Techne, SA-001), following the manufacturer’s instructions. Here, the 12–230 kDa Jess/Wes Separation Module was used. Briefly, the kit provides five SARS-CoV-2 recombinant viral antigens: RBD, N, S1 subunit, S2 subunit and S (S1+S2). The antigens were electrophoretically separated according to their molecular weight to create a ladder for capture of reactive serum antibodies. Two microlitres of plasma samples diluted 1:10 in Serum diluent buffer were loaded. For the secondary antibody, ready to use HRP-conjugated goat anti-human IgG, IgA or IgM antibody was used. Digital image of chemiluminescence of the capillary was captured with the Compass Simple Western software (version 4.1.0, Protein Simple), that automatically calculated chemiluminescence intensity of each single antigen binding signal. Results could be visualized as electropherograms representing peak of chemiluminescence intensity and as lane view from signal of chemiluminescence detected in the capillary. To control for differences in signal between experiments, a reference sample, the NIBSC Anti-SARS-CoV-2 Antibody Diagnostic Calibrant (NIBSC 20/162) was included in each experiment. A panel of pre-pandemic sera was used to calculate the negative cut-off value for each of the antigen tests (mean + 2STD). Final results of the samples were calculated by subtracting the negative cut-off value from the chemiluminescent signal of the sample.

### 2.8. Lateral flow IgG / IgM

A rapid detection kit (Accu-Tell COVID-19 IgG/IgM Antibody Test) for SARS-CoV-2 was used following manufacturer’s instructions and compared with other antibody detection platforms. Briefly, 5µl of heat-inactivated serum were added to the antigen test cassettes followed by 2 drops of the supplied PBS. After an incubation of 30 min at 20° C, the results were recorded. A positive IgG or IgM result was indicated by the appearance of a band for either of the isotypes include in the assay. Tests were valid only if a control band appeared in the device.

### 2.9. Electroporation-dependent neutralisation assay (EDNA)

Electroporation was performed using the Neon Transfection System (Thermo Fisher). Vero ACE2/TMPRSS2 cells ^35^ were washed with PBS and resuspended in Buffer R (Thermo Fisher) at a concentration of 1 × 10^6^ cells ml^-1^. For each electroporation reaction 0.5 × 10^6^ cells (10.5 µl) were mixed with 2µl of serum to be delivered. The mixture was taken up into a 10 µl Neon Pipette Tip and electroporated using the following settings: 1400V, 20ms, 2 pulses. Electroporated cells were transferred to medium supplemented with 10% serum without antibiotics. 1.5 × 10^4^ electroporated cells were seeded into 96-well plates in triplicates and after 24h transferred to containment level 3 laboratory. Supernatants were removed and all wells washed with PBS to remove any remaining antibodies that could interfere with virus entry. Cells were infected at moi = 1 in DMEM supplemented with 2% FBS and antibiotics and incubated for 24h to allow for a single replication cycle. After incubation plates, were immediately frozen at −70°C to help with cell lysis. Next, plates were thawed at 4°C and 1 volume of lysis buffer (0.25% Triton-X100, 50mM KCl, 100mM Tris-HCl pH 7.4, glycerol 40% and RNAsecure from Invitrogen at 1/100) was added to wells and mixed gently by pipetting. After 5min of lysis, cell lysates were transferred to PCR plates and virus inactivated at 95°C for 5min. RT-qPCR was performed with Luna® Universal Probe One-Step kit (E3006, NEB) following manufacturer recommendations. Primer/probe for genomic viral RNA were CDC-N2 (IDT 2019-nCoV RUO kit). Primer probe for 18S control were described previously ^36^. SARS-CoV-2_N_Positive control RNA from IDT (10006625) was used as standard for the viral genomic N reactions. For 18S standard, DNA was synthesized and kindly gifted by Jordan Clarks and James Stewart (University of Liverpool). Final concentrations of 500nM for each primer and 125nM for the probe were used. RT-qPCR reactions were run on ABI StepOnePlus PCR System (Life Technologies) with following program: 55°C for 10min, 95°C for 1min and then 40 cycles of 95°C denaturation for 10sec and 60°C extension for 30sec. RNA copy numbers were obtained from standards and then genomic copies of N normalised to 10^10^ copies of 18S. Finally, all data was normalized to 100% to PBS electroporated cells.

### 2.10. Statistical Methods

We calculated log IC_50_ values to summarise the RBD-specific and N-specific antibodies as measured by ELISA and neutralising antibody titre as measured by neutralisation. Log IC_50_ values were estimated by fitting four parameter log-logistic regression dose response curves in the R package drc ^37^. The four parameters of this curve are the minimum response, the maximum response, the log of the dilution halfway between the two (IC_50_), and the gradient at the IC_50_. Our models actually estimated the natural log of IC_50_ values because it improved model convergence and produced normally distributed values for downstream analyses.

To ensure IC_50_ values were comparable, a single gradient, minimum, and maximum value was estimated for dose response curves of all samples. To minimise noise between experimental runs the gradient, minimum, and maximum parameters were estimated based on a random subset of 200 samples and fixed for all other samples. Graphical checks showed that these parameters produced curves that fit the observed data well. We observed that this parameter fixing decreased the variance in estimated log IC_50_ values for calibrants.

Samples and calibrants could be assigned an international unitage based on their potency relative to the international standard NIBSC 20/136 which has been assigned an arbitrary unitage of 1000 IU / ml. Unitage for a sample was expressed as

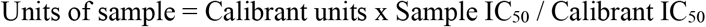

In practice, the strength of calibrants was quantified in international units as shown above and the strength of samples was calculated based on their potency relative to a calibrant with a known international unitage. The reason for this two-step process is that the international standard was not available until December 2020.

To assign international units to the calibrants, all calibrants were run in duplicate alongside the international standard and relative potencies and international units were calculated as described above. The assumption of parallel curves was verified by comparing the AIC of models which allowed separate gradients to those which did not.

The Spearman’s rank correlation coefficient for variables pairs and Mann-Whitney U tests were calculated using R (R Core Team (2019) ^38^.

## 3. Results

The primary objective of this work was to identify relevant biomarkers of humoral immunity that can serve as potential Correlates of Protection (CoP) of COVID-19. In this study we analysed antibody-based parameters present in serum of convalescent patients and compared to those in seropositive and seronegative health-care workers (HCW).

### 3.1. Clinical details of patients and healthcare workers included in this study

The participants of this study were classified into three cohorts: a) Patients; b) Seropositive HCW; and c) Seronegative HCW (Table 2). The serological status of the participants was determined by a Luminex-based assay, a commercial S and N antibody assays (Roche, ECLIA) and a commercial lateral flow diagnostic test (AccuTell). Any participant displaying a positive result by any of the screening tests was considered seropositive. A large proportion of hospitalised patients (82%) presented a clinical score of 4 (Severe Pneumonia) (Table 1b). Approximately half of these patients (47%) presented septic shock or sepsis (clinical scores of 7 and 6), 38% developed ARDS (Acute Respiratory Distress Syndrome) or severe pneumonia (clinical score 5 and 4) and only two patients presented moderate pneumonia (clinical score 3). Only two patients were asymptomatic. All patients had a positive PCR result and all, except two, presented SARS-CoV-2-specific antibodies. Clinical scores of seropositive HCW ranged between 1 and 3. A third of these individuals (33%) presented moderate pneumonia (clinical score of 3), 50% showed mild disease (clinical score 2) and 12.5% were asymptomatic. In contrast, the majority of seronegative HCW were asymptomatic (59%) or presented with symptoms of mild disease (33%) and only 3 individuals presented moderate pneumonia.

### 3.2. Calibration and standardisation of antibody assays

Whenever possible, we defined candidate humoral CoP in international units. Quantitative antibody assays (pMN, RBD ELISA, N ELISA) were calibrated using our internal reference antiserum (HICC S2) or an external calibrant. At the start of this work, the WHO International Serum, NIBSC 20/136 had not yet been developed and we calibrated our internal standard against the NIBSC 20/162 serum. In some instances, we used the latter reagent directly as our assay calibrant neutralisation (nAb) assay. This reagent was assigned 1000 units for nAb, the pan-Ig N and the pan-Ig S assay. The results of the nAb, N-ELISA and RBD-ELISA, were converted to Binding Antibody Units (BAU) once the NIBSC 20/136 became available.

In order to assign an International Unitage to our internal calibrants, which we used to determine the unitage of every sample tested by the nAb and pan-Ig ELISAs, we tested in the same assay serial dilutions of the HICC internal reference sera and NIBSC reagents. After preparing the corresponding calibration curves (Fig 1), we performed a parallel line analysis. Such analysis supported the mathematical derivation of a unitage value for our internal calibrants from the NIBSC 20/162 reagent. Thus, HICC Serum-2 was assigned a value of 504 BAU / ml for the pan Ig N-ELISA, 98 BAU / ml for the pan Ig-G RBD ELISA and 76 IU/ml for the neutralisation assay. The results of each sample tested by these assays were expressed in the corresponding units as follows:

**Figure 1:**
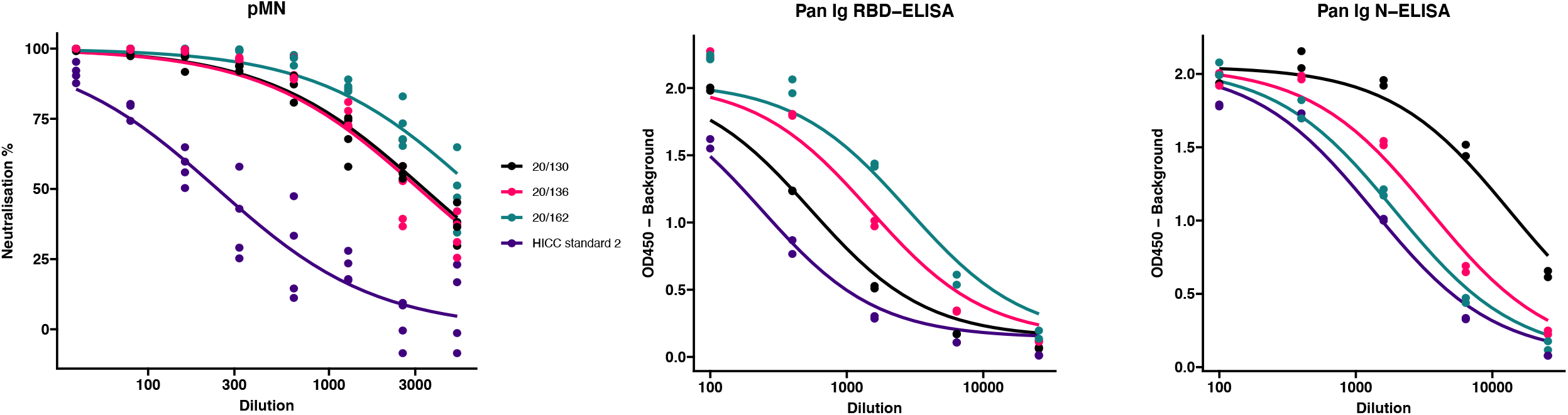
Comparison of internal calibrants and International Standard for neutralisation and binding assays. The IC_50_ from these curves were used to calculate the A) International neutralising units (IU); B) RBD-specific Binding Antibody Units (BAU); and C) N-specific Binding Antibody Units (BAU). Calibrants were run four times at each dilution for pMN and twice for ELISA tests.

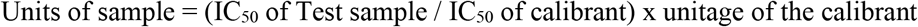

When the WHO Reference Antiserum 20/136 became available, the conversion into International Units (IU) and Antibody Binding Units (BAU) specific for N-, RBD-, and S-antigens (N-BAU, RBD-BAU, S-BAU) was calculated by multiplying the values (units) of samples by a factor F, which is the ratio of the IC_50_ of NIBSC 20/162 relative to the IC_50_ of the WHO International Serum NIBSC 20/136. Thus, all results of the neutralisation and the Pan-Ig ELISA tests included in this study are expressed in Binding Antibody Units (BAU) relative to the International Standard.

### 3.3. Antibody biomarkers of COVID-19 immunity as potential CoP

The convalescent serum samples from these three cohorts were analysed by a range of assays that measure antibody-based biomarkers of immunity: a) a pseudotype-based microneutralisation assay (pMN); b) a Luminex IgG assay specific for N, S and RBD; c) a pan-Ig ELISA for N and RBD; d) a multiplex antigen (S, S1, S2, N and RBD) immuno-blotting assay for IgG, IgM and IgA; e) a commercial lateral flow assay for rapid detection of SARS-CoV-2-specific IgG and IgM; and f) a commercial electrochemiluminescence assay (ECLIA).

We analysed the data of all these assays and determined individual correlations of all these measurements with one another and with the clinical severity scores assigned to the individuals the samples derived from. These analyses (Fig 2a) showed four main clusters of assay correlations. The first cluster is represented by IgA Immunoblotting assays for S, S1, S2 and RBD (Fig 2b). A second group is formed by IgG assays based on Spike-derived antigens and pMN assay (Fig 2b). The third cluster is formed by N-specific assays (Fig 2c). Interestingly, the intracellular neutralisation assay (EDNA) correlated positively with N-specific IgG and IgA binding assays. Not surprisingly, the IgM assays did not show positive or negative correlations with the IgG, IgA, pMN assays, intracellular neutralisation or clinical severity. Overall, clinical severity correlated positively with nAb data, S/RBD, N antibody binding measurements. As expected, nAb data correlated more strongly with S-specific and RBD-specific binding antibodies than with N-specific antibody levels, indicating N-specific antibodies maybe a good biomarker of previous infection and its severity but not necessarily the best surrogate of nAb.

**Figure 2a:**
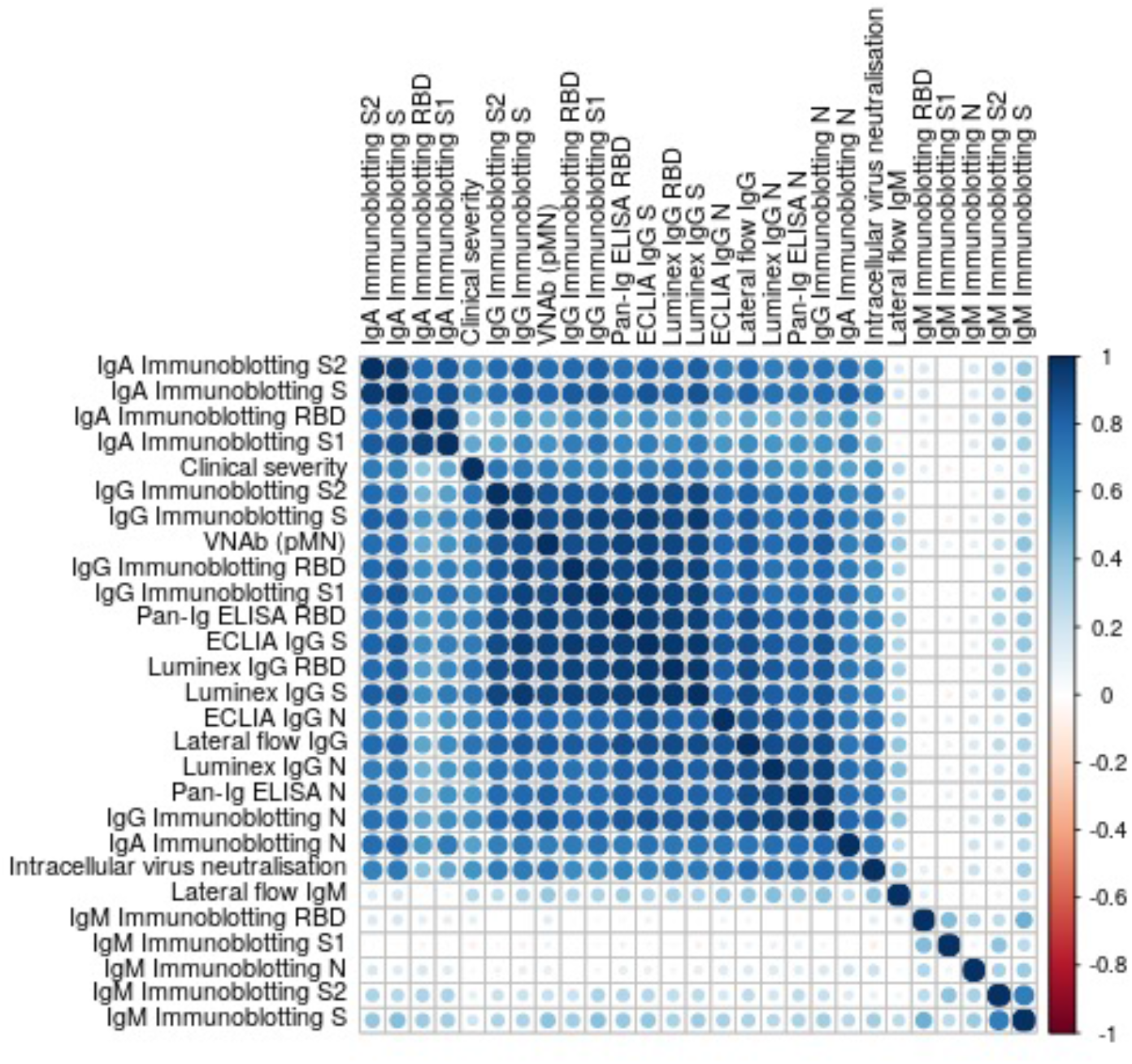
Correlation plot showing pairwise Spearman’s rank correlation between assays. Darker and larger points indicate stronger correlations. Blue indicates positive and red indicates negative correlations. Assays are ordered by hierarchical clustering so that assays with similar relationships are together.

**Figure 2b:**
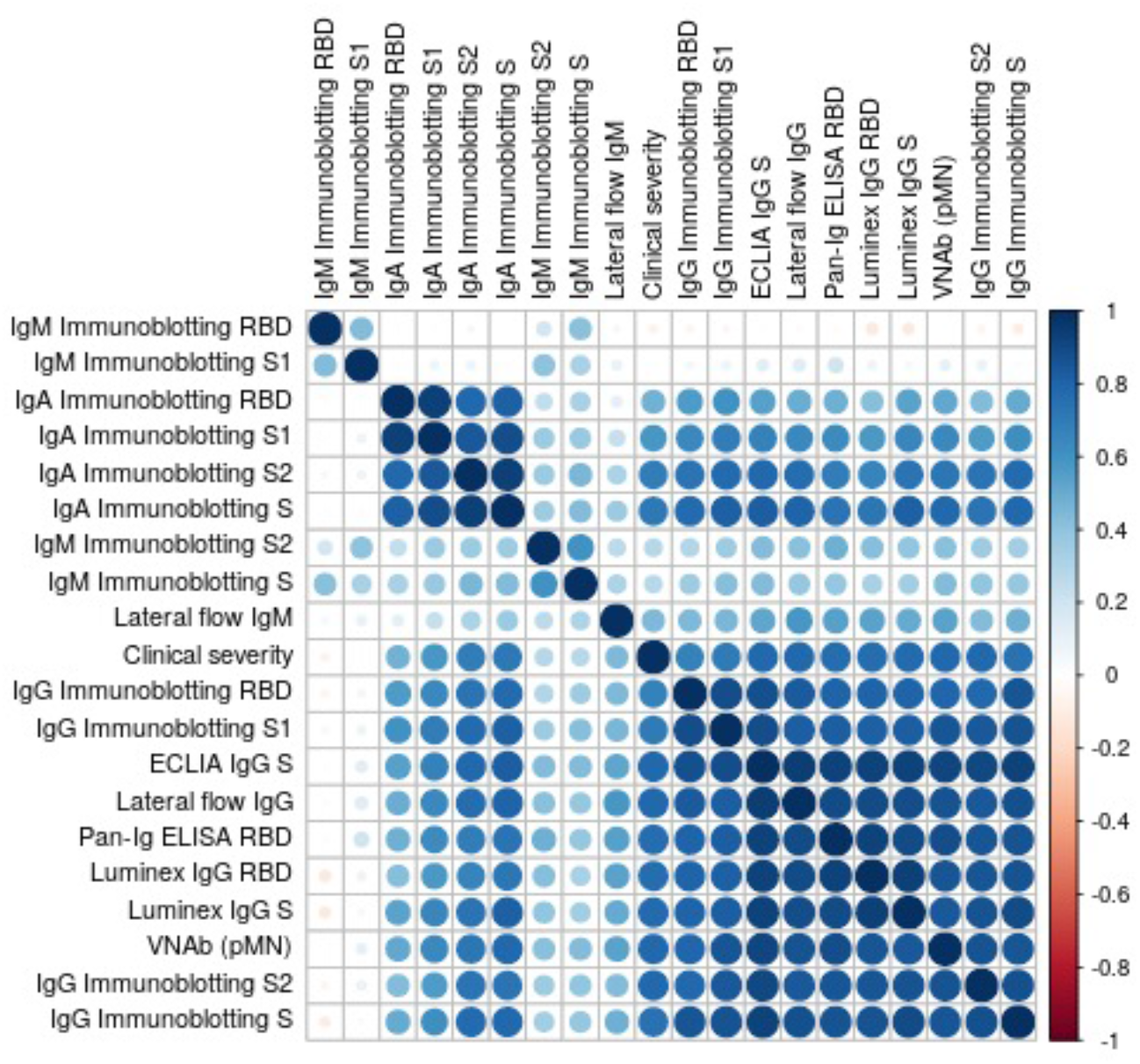
Correlation plot showing pairwise Spearman’s rank correlation between clinical severity, pMN and S-specific antibody assays. Darker and larger points indicate stronger correlations. Blue indicates positive and red indicates negative correlations. Assays are ordered by hierarchical clustering so that assays with similar relationships are together.

**Figure 2c:**
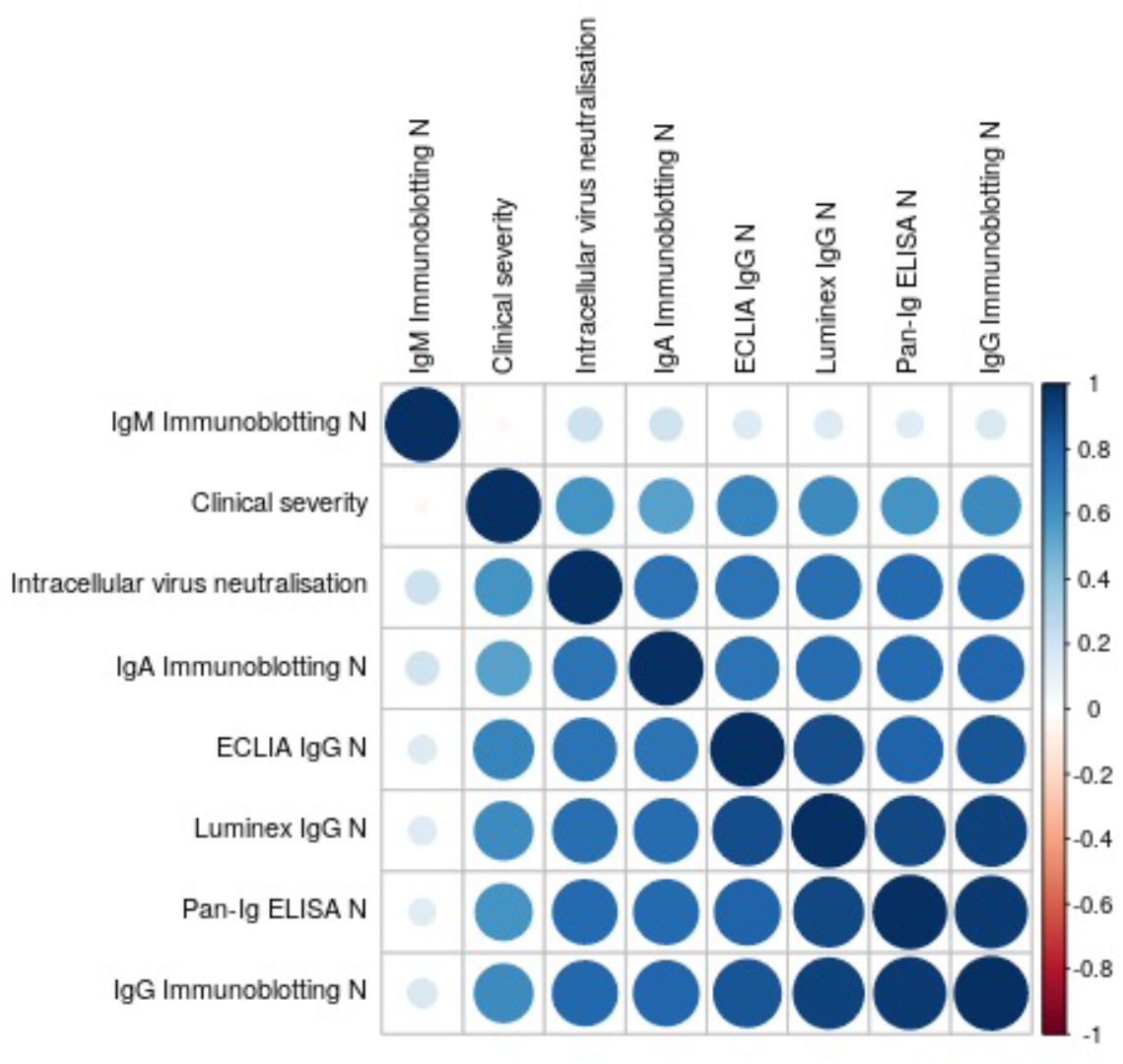
Correlation plot showing pairwise Spearman’s rank correlation between clinical severity, intracellular neutralization and N-specific antibody assays. Darker and larger points indicate stronger correlations. Blue indicates positive and red indicates negative correlations. Assays are ordered by hierarchical clustering so that assays with similar relationships are together.

Having established the general correlations of the biomarkers under study in these convalescent samples, we dissected in more detail the data generated by these assays.

### 3.4. SARS-CoV-2 antigen specific antibody responses

The antibody screening assays used to classify the serum samples from patients and HCW produced concordant results with a few exceptions. All samples from patients were positive by all three IgG assays except those from patient 37 and patient 50, which tested negative by all three assays. These patients were asymptomatic and their clinical histories revealed that they were already hospitalised for other conditions before becoming infected with SARS-CoV-2. Only a small group of seropositive HCW samples produced discordant results. Thus, Staff HCW 198, 223, 286, 370, 398 and 423 presented positive results only by some of the assays but none of these results were strongly positive. As expected, a proportion of the convalescent samples from patients and HCW did not test positive by the IgM lateral flow assay (data not shown).

The serum samples classified into the three cohorts by the screening assays were analysed by the ‘HICC in-house’ pan-Ig ELISA for RBD and N antigens. The results were largely consistent with those of the Luminex, lateral flow assay and ECLIA tests. Only a few discrepancies were noted. Thus, all sero-negative HCW samples tested negative by the N and RBD pan-Ig ELISA, except Staff 195, Staff 296 and Staff 269 samples. HCW samples Staff195 and Staff196 were positive for RBD, presenting values of 4.4 and 2.5 RBD-BAU /ml respectively, just above the negative cut-off value (2 RBD-BAU / ml), but were negative by the N ELISA (negative cut-off value of 4 N-BAU /ml). Staff 269 which had 14.3 N-BAU / ml (negative cut-off value 7.7 N-BAU / ml) but was negative for RBD. As expected, all patients’ samples tested positive against both antigens and presented high values (mean 414.5 RBD-BAU / ml; mean 316 N-BAU / ml), except patient 37, which tested negative for both antigens. Patient 50, which tested negative by all screening serological assays, presented low antibodies to RBD (20.9 N-BAU / ml) and N (26.3 N-BAU / ml) antigens. Results of the pan-Ig N and pan-Ig RBD from seropositive HCW were also in line with the Luminex, lateral flow and ECLIA tests. The average antibody levels of seropositive HCW, 118.9 RBD-BAU / ml and 234.6 N-BAU / ml, were significantly lower than those found in patients (Mann-Whitney U test, RBD: U=717, p<0.001; N U=630, p=0.006). The same samples that produced conflicting results by the serological screening assays, namely HCW Staff 198, 223, 286, 370, 398 and 423, produced very low N- and RBD-BAU results.

### 3.5. Virus Neutralising antibody responses measured by pseudotype based micro-neutralisation

The pMN results revealed a significant difference in neutralising antibody titres (nAb) between the three cohorts (Figure 3). As expected, the seronegative HCW sera presented very low nAb (mean 5.3 IU / ml). In contrast, seropositive HCW presented moderately high nAb levels (mean value 379 IU / ml), whereas patients presented a three-fold higher level (1029 IU / ml). Of note is that three seronegative HCW (Staff 38.2, Staff 38.1, Staff 228) had low nAb but these were above 24.2 IU / ml (mean of negative HCW+ 2STD). This pMN value is well above 6 IU / ml, a negative cut-off value for this assay calculated from a small panel of pre-pandemic sera (mean + 2SD) suggesting that these individuals could have been exposed to SARS-CoV-2, despite antibody binding assays showing negative values for all of them. Five seropositive HCW samples (Staff 223, 286, 370, 398 and 423) had nAb below 6 IU /ml, which was consistent with the low values obtained in the serology screening tests and the pan-Ig ELISA. All of these individuals were asymptomatic or had mild disease without pneumonia, except Staff 370 who had moderate pneumonia. It would be interesting to investigate the frequency of these cases and understand the biological meaning of these results in these particular individuals.

**Figure 3:**
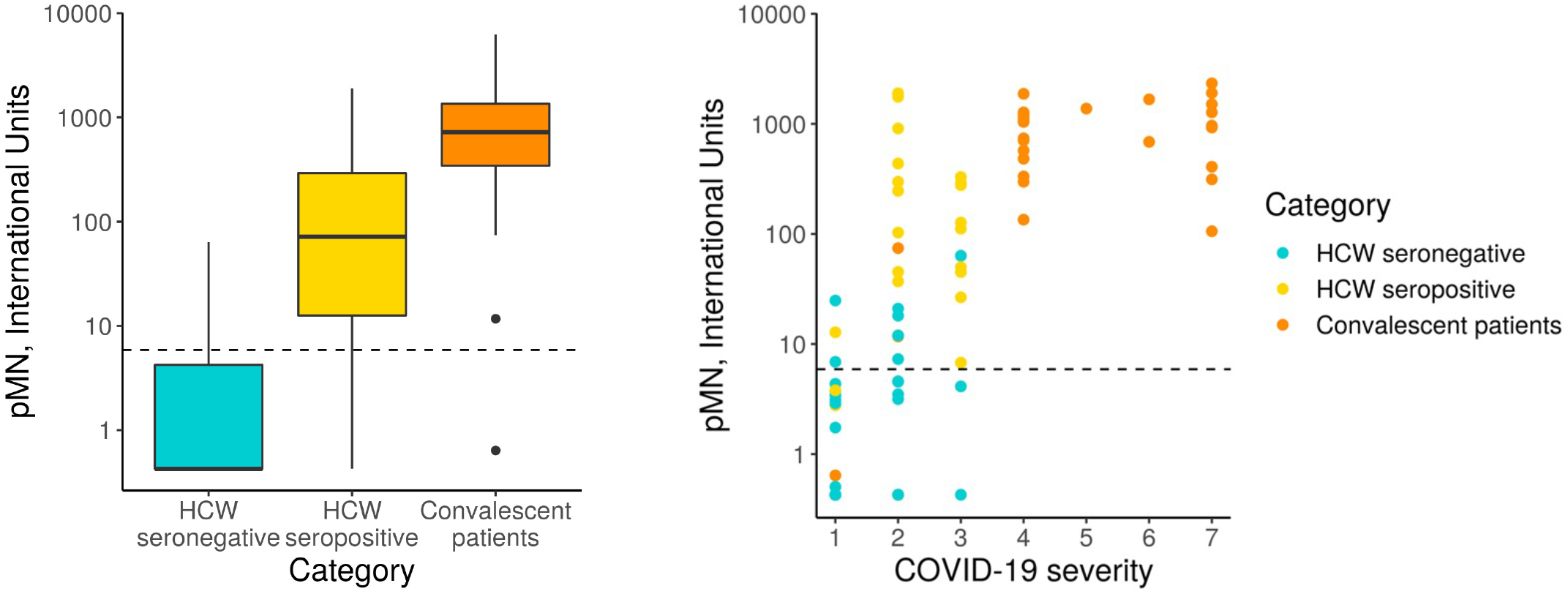
pMN virus neutralisation in International Units by cohort and COVID-19 severity. A) Boxplot showing the difference between cohorts. B) Scatterplot showing neutralisation against disease severity. The dotted line shows the 95% upper CI calculated from pre-pandemic sera, 5.9 International Units

A more detailed analysis of the data revealed a very strong correlation between clinical severity score and nAb (Figure 3, Spearman’s rank correlation = 0.71). As previously indicated in Section 3.1, the majority of hospitalised patients (clinical scores > 4) presented very high nAb levels, the majority above 200 IU / ml, and in some cases reached values as high as 2117 IU /ml. However, two patients’ samples, those from patients, 37 and 50, had levels of nAb as low as 0.63 and 11 IU /ml respectively. These samples also had low values in the ELISA and Luminex assay as discussed in the previous section. Patient 17 presented low antibody levels by both assays but remain above the positive threshold and consistent with this, presented moderate nAb levels (74 IU /ml). Interestingly, consistent with the correlation observed between severity and nAb levels, patients 17, 37 and 50 had clinical scores of 1 and 2.

The Nab data distribution in infected HCW is more widespread, ranging between 0.426 to 2092 IU / ml, including asymptomatic cases of COVID-19 (clinical score of 1) to moderate pneumonia with Sp>90% (clinical score of 3). Interestingly, some of the samples had Nab levels as high as those observed in most severe cases of COVID-19. As expected, the seronegative group of HCW presented very low nAb levels ranging between 0.426 to 18 IU /ml with clinical scores of 1 or 2.

### 3.6. SARS-CoV-2 Antigen display by automated microfluidics Western blot analysis

In order to dissect the specificity of the antibody response to Spike (S) and nucleoprotein (N) of SARS-CoV-2 and to identify additional candidate biomarkers of immunity, we used a semi-automated immunoblotting assay (Jess, Protein Simple, Biotechne) based on the separation of protein antigens in a polyacrylamide gel matrix contained in a capillary tube. This microfluidics assay sequentially processes diluted serum samples, conjugated antibodies, washing buffers and chemiluminescent reagents sequentially through microfuidics. A final chemiluminescent reaction is read by the device and translated into a luminometry intensity signal which can be analysed quantitatively. The results are visualised as traditional western blotting lane format or analytically the data outputs as densitometric units for quantitative antigen specific responses. We utilised this assay qualitatively (immuno-blotting images) and quantitatively (using total luminometry units) to screen and confirm antibody specific responses to the intact Spike protein and its subunits, S1, S2, and RBD as well as the SARS-CoV-2 N antigens. Furthermore, antibody isotype (IgM, IgG, and IgA) and IgG subtype responses (IgG1 to 4) were measured.

#### 3.6.1. Quantitative data

The results of IgG responses (Fig 4) were consistent with the findings described above for the antigen binding assays (Luminex, Lateral Flow, ECLIA and ELISA assays). The IgG antibody responses of COVID-19 patients showed significantly higher median Chemiluminescent Intensity Units (CIU) values than those of seropositive HCWs. Of note is the wide range of N-specific and S-specific IgG CI measurements, of both patients and seropositive HCW.

**Figure 4:**
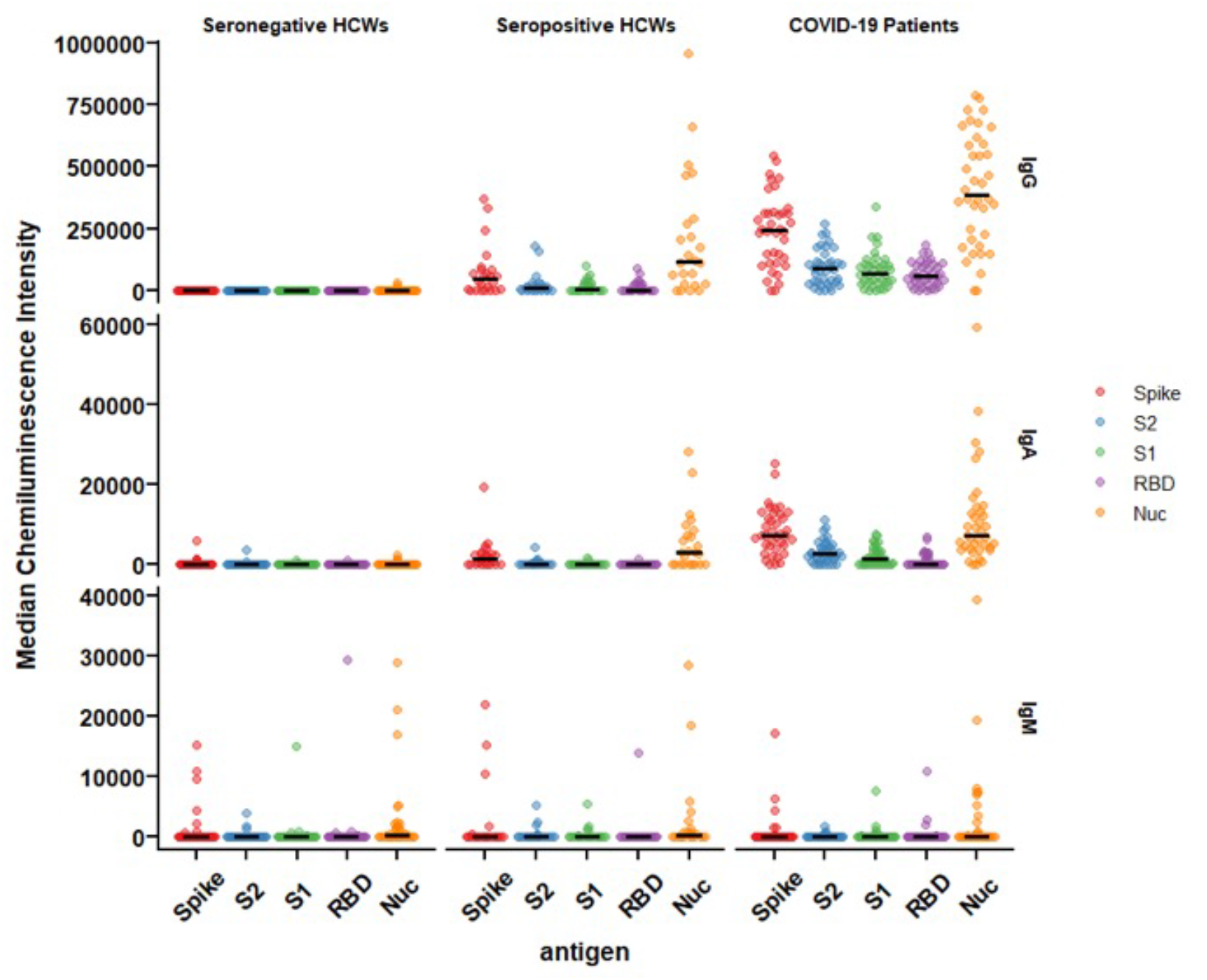
IgG, IgA and IgM responses against Spike, RBD, S1, S2 and N antigens of SARS-CoV-2. The image displays the Median Chemiluminescence Intensities of antigen-specific IgG (top), IgA (middle) and IgM (bottom) of seronegative HCWs (left), seropositive HCWs (middle) and COVID-19 patients (right). A panel of pre-pandemic sera was used to calculate the negative cut-off value for each antigen (mean + 2STD) and then subtracted from the chemiluminescent signal of each sample.

For the most part, the results of the IgA antibody immuno-blotting mirrored those of the IgG responses although the median CIU was significantly lower than for IgG, with values below 10,000 CIU as opposed to the IgG values for the same antigen in the range of 300,000 CIU. The IgA responses to the Spike subunits, RBD, S1 and S2 of seropositive HCWs against N and Spike were markedly lower than those exhibited by the IgG responses while not surprisingly, IgM responses were heterogeneous in this cross-sectional convalescent study, presenting negative or close to ‘0’ median CIU values for all three cohorts against all five antigens. Although the time of sampling was approximately 3-5 months following exposure or hospitalisation, IgM was clearly detected in a very few individuals.

#### 3.6.2. Qualitative data – Antigenic specificity of the Antibody response

Analysis of the immuno-blotting electropherograms revealed that the relative antigen response of individual sera was heterogeneous across patients and seropositive HCW revealing four distinct patterns of antigen specific responses (Figure 5). Most patients’ samples showed equally strong IgG reactivities against both N and S antigens (High N=S). However, a number of samples displayed weaker signals against both antigens (patients p17, p37, p50) (Low N=S), whereas some presented predominantly an anti-N reactivity (p64/2) (N>S) and some had predominantly anti-S-specific antibodies (p61) (N<S) (Fig 3b). This heterogeneity of the antigen specificity of the IgG response was also evident in the results of seropositive HCW samples. Again, these four categories could be distinguished according to N/S ratios: a) N = S (high) (HCWS s308, s381); b) N > S (HCWs s24, s25, s38.1, s38.2, s117, s224, s295, s361, s414); c) N < S (HCWs s249, s408) and d) N = S (low) (HCWs s4, s198, s254, s286, s294, s370, s418, s423, s439, s457). Similarly, these patterns were also identified in the IgA responses, although uniformly lower than the IgG responses in all individuals, especially in the seropositive HCW.

**Figure 5:**
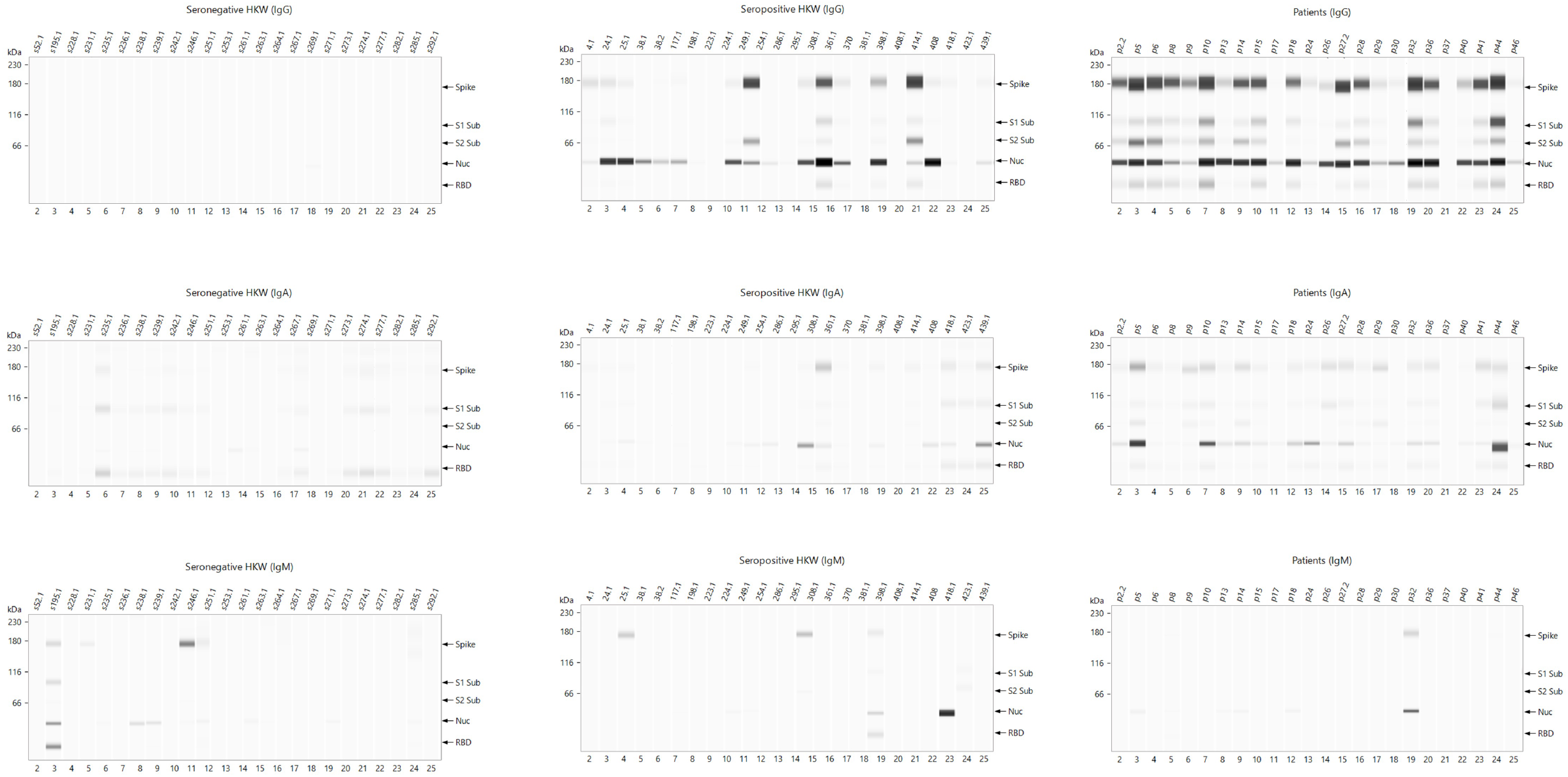
Representative electropherograms of IgG, IgA and IgM antibody responses to Spike, RBD, S1, S2 and N antigens of SARS-CoV-2 in Health Care Workers and Patients. The figure shows antigen-specific IgG (a), IgA (b) and IgM (c) antibody reactivities of seronegative HCWs (left panel), seropositive HCWs (middle panel), and patients (right panel).

There were very few samples giving a positive result in the IgM immuno-blotting assay and this signal was very low in magnitude except for one sample in each of the cohorts. For this reason the antigenic specificity patterns of the IgM response differed significantly from the IgG and IgA responses. Most positive IgM samples in the patient cohort were N-specific (n=7) and only on of the samples also had an S-specific IgM signal. Some of the seropositive HCW samples detected the Spike (HCW 25, 308 and 398) and its subunits RBD, S1, S2 (HCW 398) as well as N (HCW 398 and 418). Only samples s195 were positive for all antigens except S2, and s246, positive for S gave a clear positive signal, whereas s238 and s239 reacted to the N antigen albeit with a very weak signal. with some that had retained an exclusive N response (HCW s418). These results suggest a recent low level exposure to SARS-CoV-2 or common cold coronaviruses occurred in these individuals which was not detected by the primary screening serological tests used in this study.

#### 3.7 Intracellular neutralisation assay (EDNA)

To explore the potential use of biomarkers indicative of TRIM-21 based mechanisms of immunity we applied the EDNA assay to our cohorts’ samples. We electroporated Vero ACE2/TMPRSS2 cells with sera from each patient and seropositive HCW and ten seronegative HCW samples for reference. In the presence of electroporated N-binding antibodies, virus replication was inhibited. The results of these analyses (Fig 6) indicate that patients’ sera are more effective at inhibiting virus replication (Fig 6a), than sera from the seropositive HCW (Fig 6b). Tested seronegative HCW did not affect virus replication, as expected (Fig 5c). Interestingly, patients and seropositive HCW samples with the strongest inhibition of virus replication had the highest levels of anti-N antibodies such as staff 414 or patient 32, confirming that the observed intracellular neutralisation is mediated by anti-N antibodies. Importantly, these results highlight that traditional neutralization assays, fail to measure the potential contribution of anti-N antibodies present in SARS-CoV-2 positive sera.

**Figure 6:**
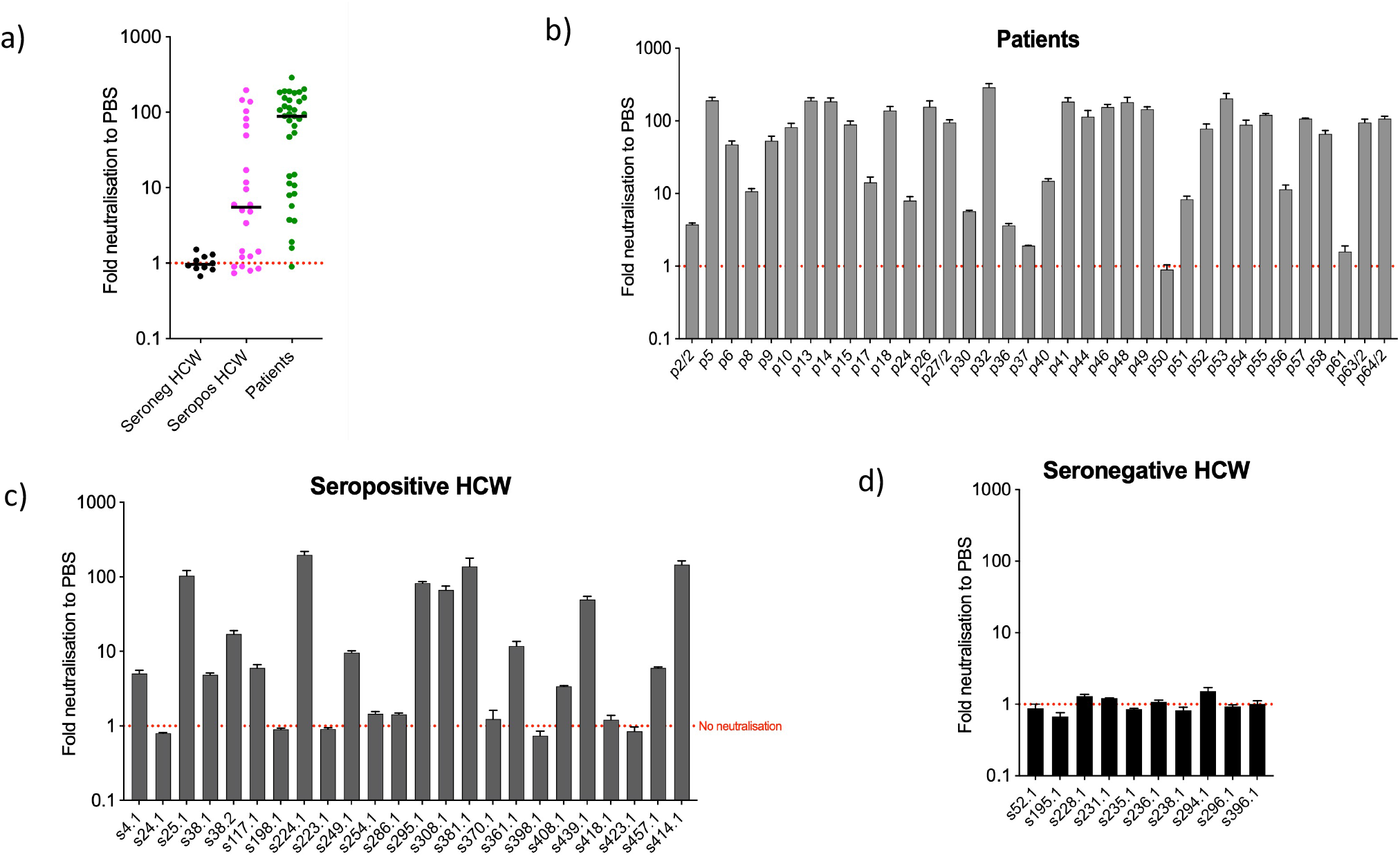
Intracellular neutralisation data from EDNA assay. The results are expressed in genome copies relative to 18S and percentage normalised to PBS. Panel a) depicts median values of EDNA results of the three cohorts, expressed as Fold neutralisation relative to PBS; Panels b), c) and d) correspond to individual EDNA results of patients, seropositive HCW and seronegative HCW respectively.

## 4. Discussion

The ‘Humoral Immune Correlates of COVID-19 Project (HICC)’ (https://www.hicc-consortium.com/) was established to identify humoral biomarkers of immunity and develop standardised assays to determine the thresholds of antibody responses that correlated with protection against SARS-CoV-2 infection or severe COVID-19 disease requiring hospitalisation. Our first objective was to define those candidate antibody biomarkers by cross-platform comparison of a range of antibody-based assays, such as nAb and SARS-CoV-2 antigen-specific (N-, S-, S1- S2- N- and RBD) binding antibodies (Pan-Ig, IgG, IgM, IgA) in convalescent serum or plasma samples from COVID-19 hospitalised patients and seropositive and seronegative HCW. The main findings confirmed: a) that there is a strong positive correlation between clinical severity and SARS-CoV-2-specific antibodies; b) that there is a strong correlation between nAb and S- and RBD-specific antibody levels; c) that intracellular neutralisation correlated very well with N-specific antibody levels; and d) that there are different antigen-specific reactivity patterns of IgG, IgA and IgM in seropositive samples. We used the WHO International Standard (NIBSC 20/136) to quantify some of these antibody-based parameters in International Units (IU) for neutralisation assays and Binding Antibody Units (BAU) for ELISA. The adoption of common results reporting unitage in IU and BAU as those described in this study would eventually facilitate comparative analyses of data generated by immunogenicity studies performed by different teams in different parts of world.

The criteria for the serological classification of the convalescent samples used in this study were based on a UKAS-accredited Luminex assay benchmarked for COVID serological screening ^32^ and another two well-stablished serological assays (AccuTell lateral flow IgG/IgM; Roche ECLIA) ^29,39,40^. The results of these and other additional tests (pan-Ig N and RBD ELISA; immuno-blotting) described in this paper were consistent cross-validated our findings. As expected, only samples with low antibody levels produced some discrepancies due to the positive-negative cut-off of each particular assay. Such discrepancies could also be due to the previously described cross-reactivity between the N antigens of SARS-CoV-2 and the seasonal human common cold coronaviruses ^41^ which was also consistent with our analysis of pre-pandemic serum samples.

Consistent with published data ^8,11,42,43^, we found a very strong correlation between nAb, as measured by the pMN assay, and severity of disease but evidence from epidemiological studies and vaccine clinical trials indicated that nAb correlate with immunity against COVID-19 ^44,45^. However, knowledge of the early immunopathologic events which trigger severe COVID-19 disease is still incomplete and the role of complement and the interaction with early antibody responses are under investigation.

There is accumulating data that supports the observation that the high nAb titres in severe COVID-19 patients are a consequence of the high and persistent viral replication, high virus load and the marked expression of viral antigens in the host or a consequence of a disregulated immune response leading to antibody-mediated immunopathology. Studies by Garcia-Beltran and co-workers suggested the latter and that the antibody response profile in severe patients, characterised by high nAb to IgG-RBD ratios, is a consequence of such disregulation. These authors suggested that the use of specific antibody response metrics could be useful to discriminate between immuno-pathological antibody responses from those that would lead to protective immunity. Our study does not address this question but proposes different antibody-based assays, parameters and standardised methods that could facilitate comparative data analysis of humoral immunity. Use of alternative antibody response metrics using these parameters could be applied in longitudinal studies of the antibody response to SARS-CoV-2 in order to elucidate thresholds of protective immunity.

Often, COVID-19 serological studies published to date show a positive correlation between Spike-specific antibodies and nAb ^46^. Accordingly, our study showed that Nab of COVID-19 convalescent sera correlated very strongly with Spike-specific IgG and IgA binding antibodies. This can be potentially very advantageous for assessing protective immunity in clinical trials or in immuno-surveillance programmes, as evidence supporting the use of nAb as a biomarker of COVID-19 immunity continues to grow ^47^. Indeed, some of the S-specific antibody binding assays used in our study are quantitative, reproducible, suitable for calibration to the international standard and high-throughput. The latter is a distinct advantage over the more laborious and time-consuming neutralisation assays. However, validity of these correlations need further evaluation as other reports indicate the importance of IgM and IgA contribution to virus neutralisation, and that the nAb / IgG ratio correlate with 30-day survival ^11^.

Our data also showed a strong correlation between nAb, disease severity and N-specific IgG and IgA antibody levels in convalescent samples. The intracellular neutralisation data generated by the EDNA assay represents an indirect evidence that the TRIM-21 mediated mechanism of immunity could play a relevant role in protection against COVID-19. The output of the EDNA assay from SARS-CoV-2 infected cells previously electroporated with serum from patients or HCW showed a significant reduction of virus replication which was proportional to the N-specific antibody levels of the sera. Previously, it has been shown that antibody-antigen complexes are rapidly degraded in the cytosol by TRIM21 and the proteasome ^17,48^. If N protein is degraded inside an antigen presenting cell, this provides peptides for MHC-I presentation. Indeed, studies have shown that cytotoxic T-cell immunity to virally-infected cells requires internalization and cross-presentation of virus-antibody complexes by dendritic cells ^49^. It has been previously shown that TRIM21 uses anti-N antibodies to degrade the nucleoprotein of LCMV, promote cytotoxic T-cells and clear mice of infection ^18^. As indicated earlier, longitudinal analyses of sera from patients and HCW will help to determine the relevance of this anti-SARS-CoV-2 immunity mechanism and how N-specific antibodies contribute to immunity or immune-pathogenicity of COVID-19. But here we provided experimental evidence that intracellular virus neutralisation is higher in patients than in seropositive samples, that it is absent in seronegative sera, and that these measurements correlate very strongly with anti-N antibody levels (Fig2).

The analysis of the antigen specificity of serum IgG and IgA of patients and HCW showed an overall immuno-dominance of N- and S-specific antibodies over S1, S2 and RBD antigens. However, we observed, consistent with other studies ^50^, that the N/S ratios were not always homogeneous as described in the results section. Further analyses of the evolution of antigen-specific antibody responses of our cohorts over time will help to interpret the relationships between these metrics and the clinical outcome of SARS-CoV-2 infection.

Our data indicate that serum IgA responses paralleled those of IgG in terms of antigen specificity, albeit the magnitude is significantly lower. Mucosal IgA might represent a critical component of the immune response against COVID-19 ^51,52^ as it contributes to virus neutralisation ^53^. Interestingly, studies have stablished a correlation between serum IgA levels and severity, with mild COVID-19 cases, such as those occurring in the young, showing secretory IgA responses with little detection of IgA in serum ^54^. In our study we have found clearly IgA levels in convalescent samples collected 3-5 months post-infection but consistent with Sterlin’s findings ^53^ the levels are significantly reduced in comparison with IgG titres. However, Varadhachary and co-workers ^52^ have detected peak IgA levels in saliva at 3 months post-infection suggesting the kinetics of IgA in serum and mucosal surfaces are different. In our study we did measure mucosal IgA and we were unable to determine the level of correlation between serum and mucosal IgA but further efforts should be aimed at elucidating how these two isotypes evolve in time in different body compartments in order to define an IgA-based biomarker of protection.

After a viral infection, IgM responses are usually the first to appear in serum n and this is the case too for COVID-19 ^55,56^. Our data indicates that IgM are easily detected in a number of individuals from the patients and seropositive HCW cohorts. Some studies report IgM lasting up to at least 3 months post-infection ^55,57^ and it is therefore not surprising that IgM is detected in some of our convalescent samples.

A cornerstone of this study was the use of the WHO standard to benchmark neutralising antibody responses and to relate other binding assays to our findings with this standard. We expressed in International Units (IU) and Binding Antibody Units (BAU) the results of the most commonly used serological assays. For this, we used the International Standard Serum 20/136, as recommended by the WHO ^24^. The objective of this approach is to develop an internationally accepted unitage to quantify the levels of cardinal serological (antibody) biomarkers of COVID-19 immunity in order to facilitate cross-comparison of immunogenicity data, which ultimately will facilitate the derivation of Correlates of Protection against COVID-19. This may become increasingly important for bio-regulatory approval of SARS-CoV-2 vaccines in future months. Indeed, the emergence and rapid spread across the globe of COVID-19, prompted the rapid development of vaccines against this disease. However, the vast amount of scientific data arising from clinical trials and epidemiological studies addressing COVID-19 immunity have not yet translated into the unequivocal definition of a reliable CoP. Vaccines that are now being used across the globe were licensed on the basis of vaccine efficacy data obtained in placebo controlled clinical trials. These are very costly and they depend on the rates of natural infections occurring in the populations to which the vaccinated participants belong. However, more vaccines are needed to meet the global public health demands, even more so with the emergence of SARS-CoV-2 variants of concern with proven ability to escape the antibody responses developed against vaccines or previous infections (https://www.sciencedirect.com/science/article/pii/S0092867421002981). But the exposure of participants to natural infections in placebo-controlled clinical trials, are increasingly difficult to justify. Furthermore, recruitment of seronegative volunteers will become more and more complicated with the continuing rise of SARS-CoV-2 seroprevalence in the global human population. In these circumstances, non-inferiority clinical trial designs and immune-bridging using an existing vaccine as a comparator would be favoured. The definition of a CoP in International Units would help assess clinical efficacy of COVID-19 vaccines on the basis of analyses of immunogenicity data, rather than relying on evaluating clinical efficacy. Recent studies point to Nab as a reliable indicator of vaccine induced immunity ^47^. However, the majority of these studies used disparate assays and units to define antibody protection thresholds. The use of an International Unit for this purpose would enable comparative analyses of immunogenicity data to be made facilitating the derivation of CoP.

In conclusion, we have identified a range of assays and biomarkers of COVID-19 immunity that will be used to define CoP in future studies using serum and plasma samples sequentially collected from these or similar cohorts to those studied here. Such studies would need to extend their focus to SARS-COV-2 variants of concern that have been emerging since the beginning of the pandemic. The emergence of these strains with enhanced transmissibility, pathogenicity and antigenicity represents another challenge for vaccine manufacturers and regulators, and developing methods for standardising assays for comparison of Nab against VOC should be a priority.

## Data Availability

All data is available referred to in the manuscript is available on request

## 5. Acknowledgements

This study was undertaken by the Humoral Immune Correlates to COVID-19 (HICC) consortium, funded by the UKRI and NIHR; grant number G107217 (COV0170 - HICC: Humoral Immune Correlates for COVID-19). RW received funding from the StMWK (ForCOVID, Bavaria, Germany). We thank the RPH Foundation Trust COVID-19 Research and Clinical teams for supporting recruitment to this study, HCWs and Outpatients who participated in this study.

